# Virtual Reality applied to Post-Stroke Rehabilitation: design and development of the NeuroRehab VR Software

**DOI:** 10.64898/2025.12.17.25342104

**Authors:** Mercedes Gil-Rodríguez, Laura Amaya-Pascasio, Alba Hernández-Martínez, Marta Rodríguez-Camacho, Manuel Fernandez-Escabias, Sofia Carrilho-Candeias, Mariam Ramos-Teodoro, Andrea Rodriguez-Solana, Andrea Orellana-Jaen, Rodrigo Fernández-Escabias, María Tomás-García, Belén Castro-Ropero, Laura del Olmo-Iruela, María Isabel López López, Karol García-Luna, Fernando Morales-Márquez, Laura Garrigo González-Garzón, Silvia Gómez-García, Irene Pérez-Ortega, María del Mar Álvarez Ariza, Mónica Rodríguez Pérez, Antonio Rodríguez-Sánchez, Inmaculada Villegas-Rodríguez, Francisco J. Amaro-Gahete, Alberto Soriano-Maldonado, Patricia Martínez-Sánchez

## Abstract

**Objective:** To describe the design and development of NeuroRehab VR, a fully immersive, specific and gamified virtual reality (VR) software aimed at improving the quality of life and reducing disability in post-stroke patients.

**Methods:** A public-private collaborative research project was carried out between 2022 and 2024 by a multidisciplinary and multicenter team comprising neurologists, rehabilitation specialists, physiotherapists, exercise and sport sciences professionals and members of the company Dynamics VR Rehab, including engineers, developers, computer programmers, game designers, and digital artists. The project was structured into three phases: preproduction, production, and postproduction, with periodic focus group meetings and testing sessions with patients in the subacute phase of stroke held every one to two months.

**Results:** In the Preproduction phase, the multidisciplinary team discussed the initial concepts and, using the SCRUM methodology together with feedback from pilot patients, developed the software design. In this process, three thematic environments (i.e., home, nature, and science fiction) were established, along with five activity types targeting upper limb rehabilitation: fine motor skills, gross motor skills, balance, rhythmic movements, and movement speed. The software incorporated fully immersive VR, advanced hand tracking technology, and adaptive gamification elements. During the Production phase, these components were implemented and consolidated into a functional prototype. Finally, in the post-production phase, several adjustments were made after identifying minor issues, with the aim of improving activity responsiveness and refining the user experience for both patients and clinicians.

**Conclusion:** NeuroRehab VR represents a promising tool to be integrated into post-stroke rehabilitation programs and is being tested though a clinical trial. Moreover, this public-private, multidisciplinary, and multicenter collaboration model constitutes an effective framework for the design and development of technologically driven solutions applicable to clinical rehabilitation settings.

## Introduction

Stroke is a cerebrovascular disease with a substantial social and economic impact (1). It is the second leading cause of death and the primary cause of acquired disability in adults worldwide (1,2). Up to 50% of patients with stroke require assistance from family members or healthcare professionals for activities of daily living and occupational tasks, which will affect their quality of life (2). Moreover, during the rehabilitation process, they will face multiple clinical, structural, geographical, and motivational barriers, particularly in the subacute phase, in the first three months, where early intervention has the greatest impact on functional recovery (3).

Upper limb rehabilitation presents a greatest challenge in stroke recovery, as only between 5% to 20% of patients fully regain functionality, while up 50% develop spasticity (4). Standard care typically consists of physiotherapy and occupational therapy delivered in hospital settings. Emerging technologies, such as robotics or virtual reality (VR), are driving innovation and demostrate considerable potential in the field of neurorehabilitation (5).

Virtual reality applied to neurorehabilitation has rapidly expanded in recent years, emcompasing several distinct modalities (3,6,7). Non-immersive VR enables interaction with a virtual environment while maintaining awareness of the real world, typically through a computer screen or tablet. Augmented reality superimposed virtual objects onto the real world via devices such as mobile phones or tablets. Mixed reality, unlike augmented reality, requires the use of VR glasses or headsets to allow simultaneous interaction with both real and virtual elements. Finally, fully immersive virtual reality (IVR) eliminates perception of the real world through specialized hardware and software integrated in headsets, or even dedicated immersive rooms, creating an environment perceived as reality through multisensory stimulation involving sight, sound, and touch (2,6).

Although numerous studies have demonstrated that VR can improve patient adherence to rehabilitation programs and enhance functional recovery (6), the currently available market options present important limitations. Many of the available tools are based on commercial gaming systems, such as Nintendo Switch or Xbox, which are often used for post-stroke rehabilitation. However, these devices are not specifically designed for this pathology, lack a clinical rationale, and offer limited replicability across patients. The aim of the present study was to describe the design and development process of the NeuroRehab VR software (8), a fully-immersive, specific and gamified VR program intended to improve quality of life and reduce disability in post-stroke patients during the subacute phase (i.e., 3 weeks after stroke onset).

## Methods

A multidisciplinary consortium was established to develop NeuroRehab VR, a fully immersive, specific and gamified VR software for post-stroke neurorehabilitation. The project adapted an interdisciplinary, patient-centered approach focusing on individuals in the subacute phase of stroke.

The consortium was composed by: a) physical activity and sport sciences professionals from two universities (i.e., University of Almería and University of Granada, Spain); b) healthcare professionals including neurologists, rehabilitation specialists, physiotherapists, from three Spanish university hospitals (i.e., San Cecilio University Hospital (Granada), Virgen de las Nieves University Hospital (Granada), Hospital San Rafael (Granada) and Torrecárdenas University Hospital (Almería)); and c) technical professionals (engineers, developers, computer programmers, game designers, and digital artists) from the company Dynamics VR (Sevilla; https://www.dynamics-vr.es/).

The design and development of NeuroRehab VR were structured into three main phases: Preproduction (November 2022–June 2024), Production (June–September 2024), and Postproduction (ongoing).

NeuroRehab VR was developed through a public-private collaboration. The process began with a comprehensive literature review of neurorehabilitation technologies and followed an agile development strategy grounded in the SCRUM methodology, is an agile project management framework based on short, iterative work cycles called sprints. It emphasizes continuous feedback, teamwork, and adaptability, allowing progressive refinement of the product according to user and stakeholder needs. This framework facilitates iterative and incremental product development through short cycles (sprints), regular team meetings, and continuous feedback integration (Figure 1). Such an approach enabled continuous adaptation of the software design to the specific needs identified at each development stage. Regular online and in-person meetings were held with neurologists, rehabilitation physicians, physiotherapists, researchers, and technical staff from Dynamics VR.

**Figure 1.**
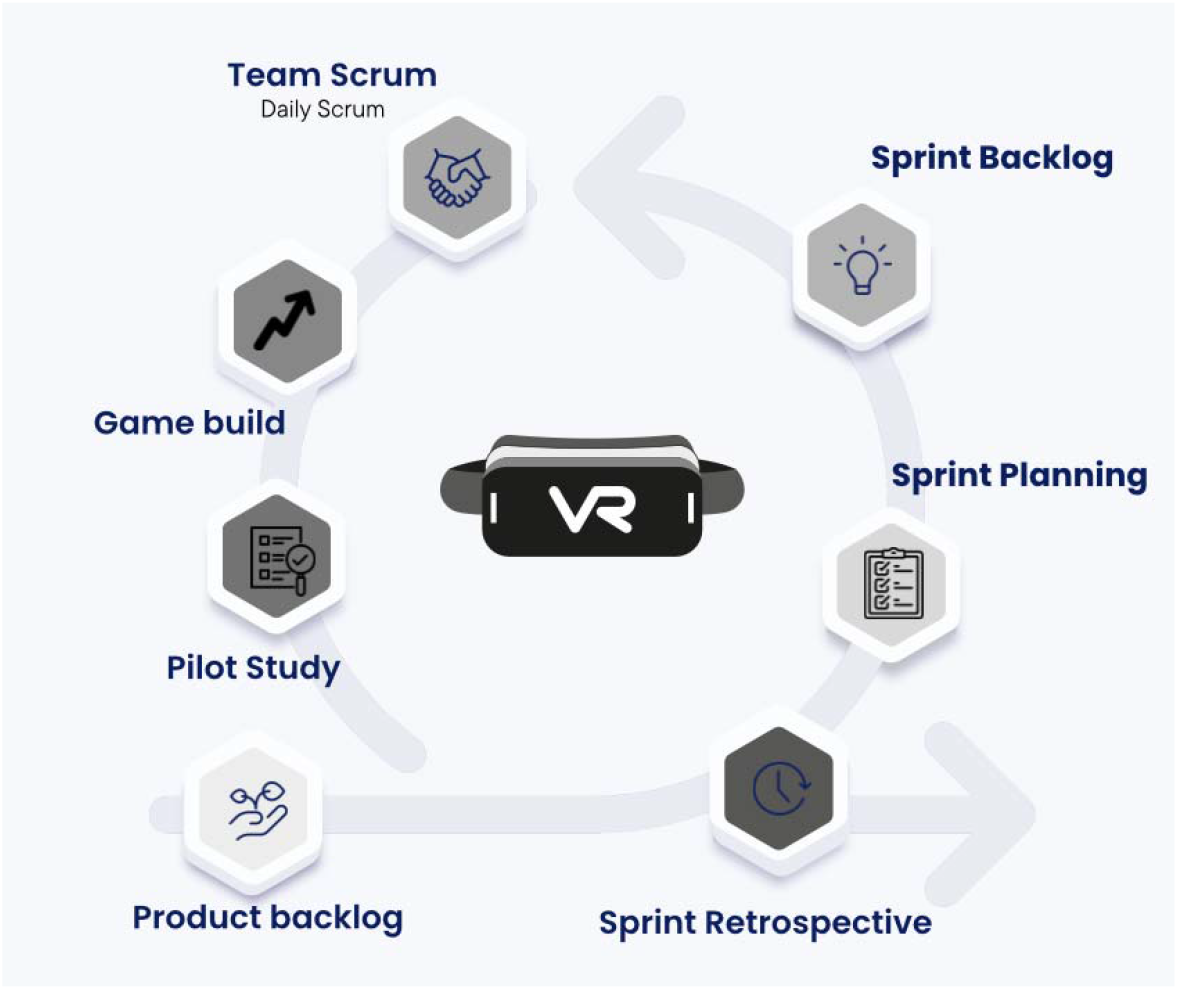
Diagram of the SCRUM methodology for the development of the Preproduction Phase. Iterative process of software design, testing, and refinement through short development cycles (sprints) and multidisciplinary team collaboration.

The NeuroRehab VR software was integrated into the META QUEST 3 headset (Meta Platforms, San Francisco, California, USA) (Figure 2) and to include specific modules targeting motor skills, balance, and cognition (8). These domains were addressed through gamified activities in virtual environments that combined everyday scenarios with imaginative tasks. Task progression was designed to occur through incremental achievements accompanied by personalized feedback to enhance motivation. The software also incorporated visual and auditory guidance to promote patient autonomy. Given that the intervention was intended for the early subacute phase of stroke (when most patients exhibit upper-limb impairments) a hand-tracking system was implemented to optimize interaction within the VR environment. Initially, the software was conceived for standing use, however, after several multidisciplinary meetings and evaluations, it was decided that all activities would be performed in a seated position to minimize fall risk and ensure patient safety.

**Figure 2.**
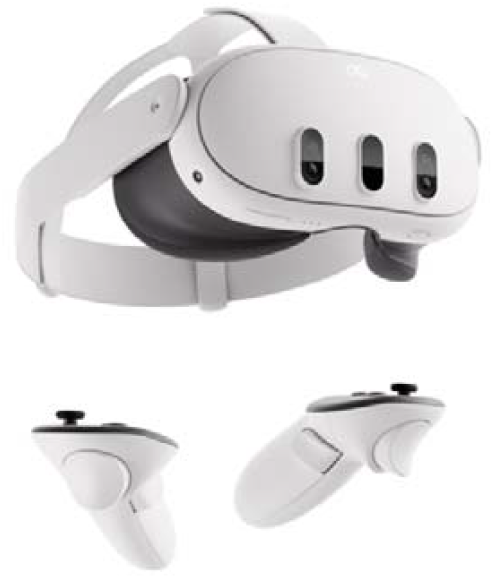
Meta Quest 3 virtual reality headset used for the design and development of the NeuroRehab VR software.

Finally, a preliminary user testing was conducted in patients with subacute stroke. Eligible participants were adults (≥18 years) with ischemic or haemorrhagic stroke occurring 7–14 days prior, functionally independent before the event (modified Ranking scale <3) (7), and presenting limb paresis, trunk stability, and adequate ability to follow instructions. Exclusion criteria include severe aphasia, cognitive or psychiatric disorders, major comorbidities, unresolved cerebrovascular conditions, or photosensitive epilepsy. Each session generated unstructured qualitative feedback collected during and after software use. This feedback was analysed by the multidisciplinary team and used to guide iterative refinements in gameplay and interface design. The continuous integration of clinical expertise, technical development, and patient input supported the optimization of user experience, therapeutic applicability, and overall feasibility.

## Results

The design and development of Neuro-Rehab VR were organized into three main phases, progressing from initial concept to clinical application (Figure 3). The key milestones of each phase are outlined below.

**Figure 3.**
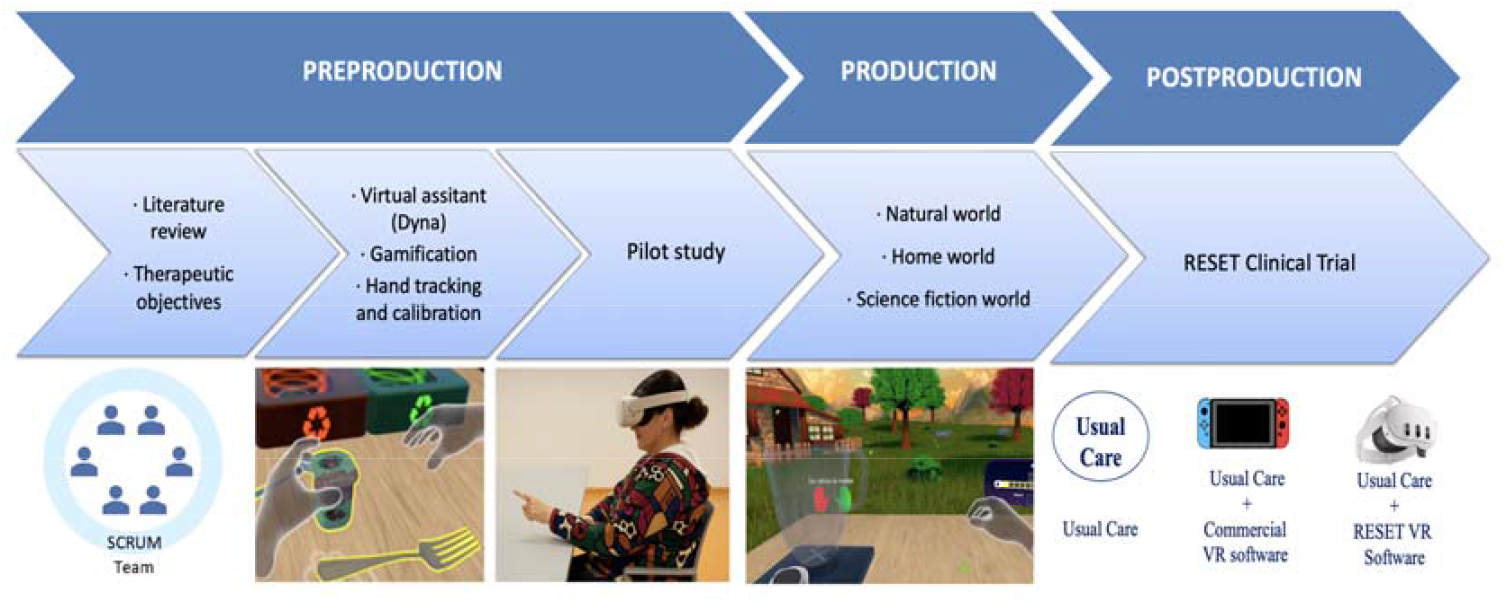
Phases of the design and development of Neuro-Rehab VR Chronological structure of the project: preproduction, production, and postproduction, from conceptualization to clinical validation. Image of a patient created using artificial intelligence.

### Preproduction phase

The process began with a literature review of major medical databases (PubMed, SCOPUS, Cochrane Library) to identify current limitations and clinical opportunities for the application immersive VR in subacute stroke rehabilitation (data not shown). Most available reports focused on non-specific commercial devices such as Nintendo or Xbox, primarily used in patients with chronic stroke (9–11). Consequently, a critical gap was identified regarding whether a fully immersive and stroke-specific VR software could further enhance the rehabilitation process in this population.

### Therapeutic objectives

Three primary goals established: (1) to increase effective rehabilitation time, (2) to promote patient autonomy, and (3) to provide a fully immersive and gamified experience.

The design of the rehabilitation activities was guided by five therapeutic domains: fine motor skills, gross motor skills, balance, movement speed, and movement sequencing. Gait-related tasks were excluded due to the risk of falls, and patients with moderate-to-severe aphasia were not included to ensure comprehension of instructions. For safety and accessibility, all activities were performed in a seated position, with a real table aligned to its virtual counterpart within the simulated environment.

**Table 1.** Summary of Preproduction Testing Results of the Neuro-Rehab VR Software.

| Thematic block | Aspect evaluated | Results |
| --- | --- | --- |
| **Therapeutic objectives** | Rehabilitation time, autonomy, immersion | Three main objectives and five priority therapeutic areas were defined |
| **Virtual assistant (Dyini)** | Need for initial guidance | Dyini, a blue butterfly, was incorporated as support during the first 4 sessions |
| **Gamification** | Motivation and adherence | Ten-level system. Ten correct repetitions required to progress to the next level |
| **Hand tracking and calibration** | Interaction accuracy | Partial embodiment removed; virtual table adjusted to real table for seated patients |
| **Preliminary user testing with patients** | Usability, complexity, and motivation | 23 patients in 5 sessions. Technical and ergonomic adjustments made |

### Virtual assistant

To facilitate user familiarization, a digital assistant named Dyni, represented as a blue butterfly, was incorporated into the software to guide users during the first four sessions. Although the initially planned yes/no interactions were not technically feasible, Dyni remained as a key orientation and support element. The first session focused on basic system interaction, while the subsequent three introduced the virtual worlds comprising the software (home, nature, and science fiction). During preliminary testing, it was observed that some patients had limited reading, writing, or computer skills. Therefore, this feature was introduced after the multidisciplinary meetings to enhance accessibility, providing visual and auditory guidance that supported autonomous learning and early engagement.

### Gamification

A progression system was implemented, requiring the achievement of three stars to complete each task and unlock the subsequent level. This adaptive mechanism automatically adjusted the level of difficulty according to the individual performance: patients who successfully executed the repetitions with the affected hand advanced to higher levels, whereas those who encountered difficulties remained at their current level improvement was achieved. Feedback was provided through visual, auditory, and written cues to reinforce comprehension, engagement, and motivation.

### Hand tracking and calibration

Hand tracking was a fundamental component of user interaction. Initial attempts at partial embodiment (arms and hands) were discontinued due to latency issues and reduced control precision. Spatial calibration represented an additional challenge, as early versions exhibited misalignments between the real and virtual tables, resulting in interaction errors. Through iterative patient testing, a precise and stable calibration process was achieved, allowing for realistic, safe, and fluid interaction within the virtual environment.

### Preliminary user testing

Five sessions were conducted with a total of 23 patients in the acute, subacute, and chronic phases of stroke. Patient feedback prompted significant technical and ergonomic adjustments. Usability, hand-tracking accuracy, task complexity, and overall immersive experience were systematically evaluated (Table 2). The iterative process, combined with direct patient input, facilitated the development of a tool specifically tailored to real clinical needs.

**Table 2.** Evolution of patient experience in the pilot meetings.

| Category | Evolution |
| --- | --- |
| **Hand tracking and calibration** | 1st: Affected hand not detected, calibration errors.<br>2nd: Clear improvement, though occasional difficulties remain.<br>3rd: Better hand recognition, but difficulty grasping some objects.<br>4th: Some calibration errors with the table.<br>5th: Minor adjustments, good detection. |
| **Instructions** | 1st: High need for guidance, poor comprehension.<br>2nd: Some patients need help, others understand activities.<br>3rd: Verbal instructions required in some cases where games are not understood.<br>4th: Verbal instructions needed for patients who cannot read.<br>5th: Game comprehension improved, but auditory support still required. |
| **Task complexity** | 1st: Games too difficult for the affected hand.<br>2nd: Some games cannot be performed with the affected hand.<br>3rd: Progressive adaptation of difficulty.<br>4th: Alternative tasks proposed due to excessive difficulty.<br>5th: Some games remain challenging, but patients feel motivated to overcome them. |
| **Motivation and acceptance** | 1st: High motivation despite technical problems.<br>2nd: Very stimulating; patients feel more focused on rehabilitation.<br>3rd: Games very appealing; patients describe them as enjoyable and perceive session time as shorter than reality.<br>4th: Patients feel capable of performing tasks independently and find them fun.<br>5th: Positive perception maintained; satisfaction with use reinforced. |

Table 2 summarizes the main domains analyzed during the five pilot meetings conducted by the multidisciplinary team. In each session, patients tested updated versions of the NeuroRehab VR software and provided qualitative feedback on their experience. The table reflects the patients’ perceptions and observations across four key categories (i.e., hand tracking, instructions, task complexity, and motivation) capturing their progressive adaptation throughout the sessions. Since feedback was collected through open discussion rather than structured questionnaires, the results represent subjective impressions rather than quantitative data.

### Production phase

During this phase, finalized versions of the software were developed. Each activity lasted three minutes: 90 seconds performed with the unaffected limb followed by 90 seconds with the affected limb, allowing motor learning with the healthy side before training the impaired extremity.

The three virtual environments were fully developed (Table 3): 1) Nature, an open-field environment featuring tasks such as fruit picking or line painting; 2) Home world—a domestic setting with daily-life activities including recycling, cleaning, or cupboard-based memory games; and 3) Science fiction—a futuristic environment involving task such as assisting a robot, interacting with drones, and performing balance challenges.

**Table 3.**
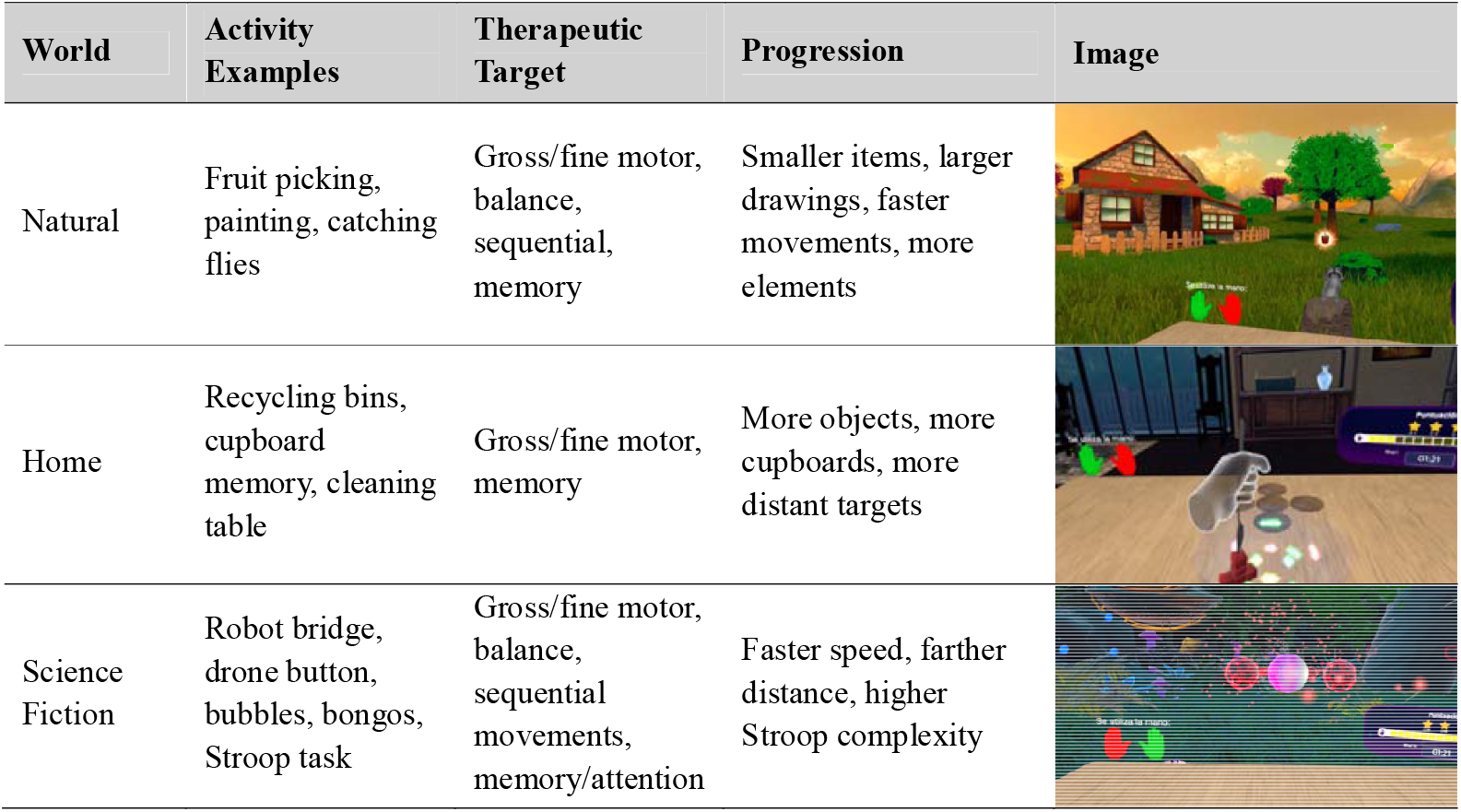
Virtual reality environments included in the NeuroRehab VR software.

| World | Activity Examples | Therapeutic Target | Progression | Image |
| --- | --- | --- | --- | --- |
| Natural | Fruit picking, painting, catching flies | Gross/fine motor, balance, sequential, memory | Smaller items, larger drawings, faster movements, more elements |  |
| Home | Recycling bins, cupboard memory, cleaning table | Gross/fine motor, memory | More objects, more cupboards, more distant targets |  |
| Science Fiction | Robot bridge, drone button, bubbles, bongos, Stroop task | Gross/fine motor, balance, sequential movements, memory/attention | Faster speed, farther distance, higher Stroop complexity |  |

Table 4 shows some examples of activities included in the three virtual environments: nature, home, and science fiction. Each environment contains five different tasks designed to train specific motor and cognitive domains. In the natural environment, the activities are Food to Plate, Magic Brush, Raccoon Challenge, Catch the Flies, and Ingredient Memory. The home environment includes Recycling, Match the Boxes, Clean the Table, Agile Basket, and Kitchen. Finally, in the science fiction environment, the activities are Help the Robot, Catch the Drone, Pop the Bubbles, Bongo Rhythm, and Color Press.

Additionally, the AppDynamics-VR platform was developed to register anonymized patients, track completed sessions, and provide clinicians with objective performance metrics. Individual results were graphically displayed in a pentagon-shape diagram, with each vertex representing one of the trained domains. This visualization facilitated rapid identification of strengths and areas requiring improvement. Patients were also able to view their own pentagon diagram, providing intuitive feedback on their rehabilitation progress.

### Postproduction phase

During the post-production stage, the development team implemented progressive adjustments to resolve minor technical issues, optimize activity responsiveness, and refine the user experience for both patients and clinicians. These improvements were carried out iteratively as the software continued to be implemented.

## Discussion

Neuro-Rehab VR represents an innovative technological approach in the current field of neurorehabilitation. Unlike other tools based on commercial video games or mobile adaptations (2,10–12), typically non-immersive systems, this application was conceived and developed from the ground up by a multidisciplinary team comprising neurologists, rehabilitation physicians, physiotherapists, sport and exercise professionals, and the technical team of Dynamics. This collaborative approach was specifically aimed at overcoming the limitations observed in non-specific devices (8,13). To date, according to the available literature, no other immersive VR software has demonstrated a comparable combination of clinical specificity and multidisciplinary integration whitin this context.

IVR was selected for software development based on the existing evidence supporting its effectiveness. For instance, the meta-analysis by Soleimani et al. (4) demonstrated the efficacy of IVR for upper limb rehabilitation in neurological patients. Additional studies have reported that active hand engagement within immersive and mixed VR environments significantly improves movement quality and functional participation of the affected limb, supports body schema integration, and reduces hemispatial neglect after stroke (14,15). Accordingly, Neuro-Rehab VR was designed as a fully immersive platform employing head-mounted displays and advanced hand-tracking technology. This setup enables patients to interact directly with the virtual environment using their own hands and arms, without external controllers. Such interaction promotes specific training of the affected upper limb, facilitates its functional integration into daily activities, and enhances engagement and motivation (15).

Although other immersive VR trials have been conducted, many involved the use of headsets and headphones for generalized activities (16) or rely on non-specific commercial systems such as Nintendo or Xbox. While these approaches have shown positive outcomes, they often lack clinical specificity or target only isolated activities (9). According to the available literature, few immersive VR platforms have been specifically developed for subacute stroke rehabilitation (2,9,10,17). Therefore, the creation of a tailored immersive VR software for this population remains a critical need.

Neuro-Rehab VR comprises three virtual environments: nature, home and science fiction, representing two familiar and one novel context. Each includes five activities aligned with therapeutic goals: fine motor skills, gross motor skills, balance, sequential movements, and memory (18). In total, fifteen gamified tasks were developed to reflect and enhance the motor and cognitive functions typically targeted in post-stroke rehabilitation. Evidence indicates that multisensory and varied activities promote neuroplasticity, support functional recovery, and improve patient engagement by reducing monotony (19,20).

The software and its tasks were refined based on feedback obtained during pilot testing, ensuring alignment with patient-specific needs. Gamification elements were incorporated to maintain engagement, while adaptive task progression provided individualized therapeutic challenges. Current evidence supports that immersive, non-immersive, and mixed VR are effective, motivating, and patient-centered interventions that enhance adherence through gamified design (21–23). Virtual assistant named Dyni, helps patients understand the initial steps and become familiar with the system, thereby promoting independence. Previous research highlights the importance of adequate initial orientation to ensure effective execution of rehabilitation tasks (24).

VR has emerged as an effective tool for upper limb rehabilitation, enhancing motor recovery, adherence, and motivation. However, most studies have focused on patients with chronic stroke, a stage in which neuroplastic potential is diminished (25–27). The Neuro-Rehab VR was specifically developed for the subacute phase of stroke, when the brain is more responsive to neuroplastic changes and cortical reorganization, thereby maximizing relearning and rehabilitation effectiveness (28). Moreover, Neuro-Rehab VR is cost-effective, operating on a commercial MetaQuest 3 headset, that is lightweight and portable compared with robotic VR systems (12). This feature enhances its feasibility in both hospital- and home-based rehabilitation, thereby improving accessibility to neurological care (29).

This study has several limitations. The software was developed and tested in a small sample of patients with subacute stroke, which may limit generalizability of the findings. Pilot testing generated qualitative rather than quantitative data. Additionally, technical issues such as hand-tracking calibration and learning demands may have influenced usability, underscoring the need for validation in larger clinical trials, including the ongoing RESET trial (8).

## Conclusions

NeuroRehab VR represents a promising tool for integration into post-stroke rehabilitation programs. Moreover, this public-private, multidisciplinary, and multicenter collaboration model offers a valuable framework for the design and development of technological solutions applicable to clinical rehabilitation settings.

## Acknowledgements

The authors would like to thank to the patients, who played an essential role in developing the software and designing this study. We also gratefully acknowledge the “Neuropsychological Assessment and Rehabilitation Center (CERNEP)”, from the University of Almería, for providing access to patients with stroke and their predisposition to help.

## DECLARATIONS

### Contributors

All authors have contributed significant intellectual content to this project. AH-M, IV-R, FJA-G, PM-S and AS-M were involved in the study design. M G-R, L A-P, A H-M and M R-C, drafted the manuscript with relevant input from M R-E S C-C, M R-T, A R-S, B C-R, L O-I, K G-L F M-M, L G G-G, S G-G, I P-O, M R-P, A R-S, and I V-R, FJ A-G, A S-M and P M-S are principal investigators, and the project administrators and guarantor and accept full responsibility for the finished work and/or the conduct of the study. All Authors have reviewed and approved the final version of this manuscript, providing written consent for publication and accepting complete responsibility for its content. The contributions of each author were agreed prior to manuscript submission.

### Patient and public involvement

Patients and/or the public were involved in the design, or conduct, or reporting, or dissemination plans of this research. Refer to the Methods section for further details.

### Patient consent for publication

Not applicable.

### Ethics approval

The study has been approved by the Human Research Ethics by the Ethics Committee for Clinical Research in the province of Almería (118/2023). Significant changes will be reported to the ethics committee. Eligible participants and their caregivers received written and oral information about the study. All participants were required to sign the informed consent.

### Provenance and peer review

Not commissioned, externally peer reviewed.

### Data availability statement

The data that support the findings are available from the corresponding author on reasonable request.

